# Capability and reliability of deep learning models to make density predictions on low dose mammograms

**DOI:** 10.1101/2024.01.01.23300313

**Authors:** Steven Squires, Alistair Mackenzie, D. Gareth Evans, Sacha J Howell, Susan M Astley

## Abstract

**Purpose:** Breast density is associated with risk of developing cancer and can be automatically estimated, using deep learning models, from digital mammograms. Our aim is to estimate the capacity and reliability of such models to estimate density from low dose mammograms taken to enable risk estimates for younger women.

**Methods:** We trained deep learning models on standard and simulated low dose mammograms. The models were then tested on a mammography data-set with paired standard and low-dose image. The effect of different factors (including age, density and dose ratio) on the differences between predictions on standard and low dose are analysed. Methods to improve performance are assessed and factors that reduce model quality are demonstrated.

**Results:** We showed that whilst many factors have no significant effect on the quality of low dose density prediction both density and breast area have an impact. For example correlation between density predictions on low and standard dose images of breasts with the largest breast area is 0.985 (0.949-0.995) while with the smallest is 0.882 (0.697-0.961). We also demonstrated that averaging across CC-MLO images and across repeatedly trained models can improve predictive performance.

**Conclusion:** Low dose mammography can be used to produce density and risk estimates that are comparable to standard dose images. Averaging across CC-MLO and across model predictions should improve this performance. Model quality is reduced when making predictions on denser and smaller breasts. Code is available at: https://github.com/stevensquires/

## 1 Introduction

Breast cancer is a leading cause of death among women below the age of 50 with women who have a known family history of breast cancer being at greater risk. Screening can offer reduced mortality [1], however, many younger women with no family history still go on to develop breast cancer [2, 3]. Assessing which younger women would benefit from early screening should enable an improvement in early detection of cancers and facilitate targeting of risk-reducing interventions.

High mammographic density is known to be strongly related to increased risk of developing breast cancer [4]. Identifying women with high breast density can thus enable preventative interventions (e.g. breast cancer risk-reducing drugs like Tamoxifen [5], which can also reduce density), additional surveillance (e.g. earlier or more frequent screening) or the use of alternative or additional screening modalities (e.g. MRI or ultra-sound scans).

There are several available methods to estimate breast density from mammograms, both automated [6–8] and by radiologists [9]. Radiologist’s time is limited and it would enable these experts to focus on more challenging problems if automated methods can be used effectively.

Younger women are not routinely screened in part because of the radiation dose risk [10]. Low dose mammography is being investigated as a means to enable density assessment in younger women which can be used to tailor their screening in the future [11].

Deep learning methods have been shown to produce mammographic density predictions with a high correlation between standard and low dose mammography [12], where the low dose images were taken at a radiation dose of around 10% of the standard dose. This suggests that using low dose mammography for risk prediction in younger women is feasible. However, more evidence is required about the relationship between density predictions on standard and low dose mammograms. One possibility for the strong correlation shown between standard and low dose mammograms is that image resolution is often reduced substantially when using deep learning methods and this reduction may obscure features that would otherwise be available in the standard dose but not low dose images. This could result in falsely similar predictions on standard and low dose images. However a previous study [13] found no evidence of this effect and proposed that the similarity of prediction on standard and low dose images does not decline at higher resolution. There are, however, many other factors that might influence the variability in prediction on standard and low dose mammograms.

In this paper we investigate where the variability in prediction occurs, how much confidence in the results we should have and how to reduce the uncertainty. Specifically if low dose mammography could be used for density, and therefore risk, estimation we need to know how closely the predictions would match predictions on standard dose images. We also need to consider how to reduce the variation between standard dose and low dose predictions.

## 2 Data

The aim of this study is to investigate how well we can estimate mammographic density in low dose (approximately 10% of standard dose) mammograms. For this purpose we utilise two data-sets which we describe in this section.

### 2.1 ALDRAM

The Automated Low Dose Risk Assessment Mammography (ALDRAM) dataset [12] was produced as part of a study to investigate whether automated mammographic density predictions could be made on images with 10% the dose of a standard mammogram. Participants were recruited from the Nightingale Centre at Wythenshawe Hospital, Manchester University NHS Foundation Trust. All participants were between 30-45, were attending for annual screening mammography, and had previously been diagnosed with unilateral breast cancer.

Right cranial-caudal (RCC), left cranial-caudal (LCC), right mediolateral oblique (RMLO) and left mediolateral oblique (LMLO) full-field digital mammograms were taken at standard dose and, whilst the breast was in position for the RCC and RMLO images, further mammograms were taken with a dose of approximately 10% of the standard dose (the low dose images). As the compression and positioning are the same, the standard and low dose images should be directly comparable. All the images are acquired using GE Senographe Essential (GE Healthcare, Buc, France) machines.

In Figure 1 we show an example of a standard dose RCC image and its low dose counterpart which have both been pre-processed from the *for processing* (or *raw*) mammograms (see Section 3.1 for details). The left two plots show examples of the whole image for the standard and low dose respectively. These are images that are fed into the density prediction model - they have been pre-processed. The dashed rectangle in the left-hand plots is a patch of size 320 × 256 which is then shown in the right-hand plots. The standard and low dose images have the same structure but the low dose image has a higher level of noise.

**Fig. 1.**
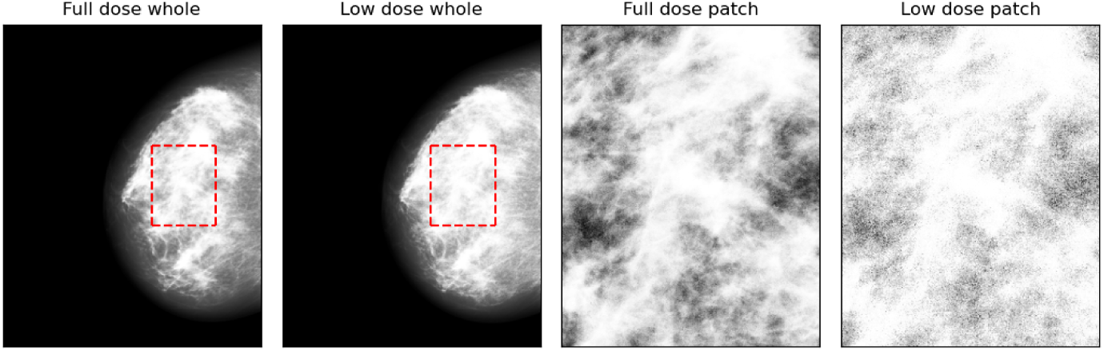
An example of a pre-processed (see Section 3.1 for details) CC image from the ALDRAM data-set with standard and low dose shown as well as two patches of the images. Left two plots) Standard dose and low dose whole images, respectively. The rectangular outline shows the location and size of the zoomed in patches. Right two plots) A zoomed in image of the outlined part of the whole images.

In total we consider 147 sets of images (the four standard dose and two low dose images). Of those image sets 82 have size (2, 294 × 1, 914) and 65 have size (3, 062 × 2, 394). Each set of images, or each participant, is made up of only one of the image sizes.

The ages of the patients range from 31 to 45 which is considerably lower than the distribution of ages in the training data-set (see Section 2.2). The age distribution is shown in Figure 2.

**Fig. 2.**
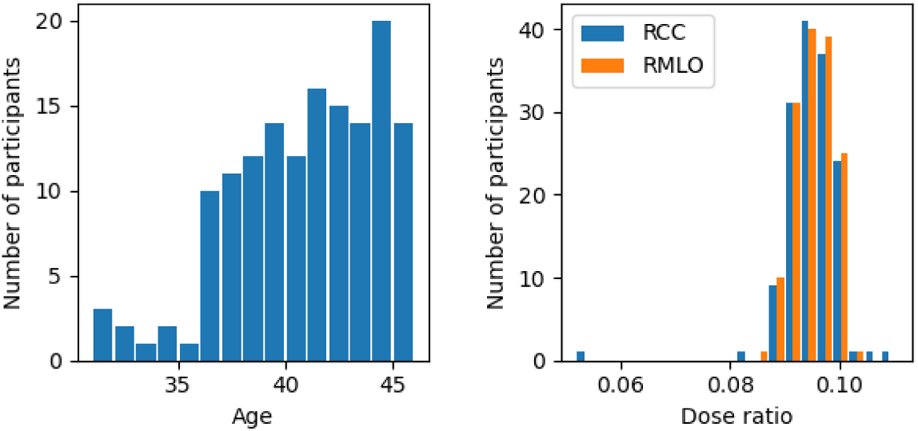
Left) Histogram of the age distribution of the 147 women in the ALDRAM dataset. Right) Histograms of the ratios of low to standard doses for the RCC and RMLO images.

The distribution of the exposure ratios between the low and standard dose for the two mammographic projections of the right breast is also shown in Figure 2. There is one outlier on the RCC side with an exposure ratio of 0.049 compared to around 0.1 for the rest. Otherwise the exposure ratios are fairly similar. There is no relationship between the exposure ratio for the RCC and RMLO for pairs of the same patient.

### 2.2 PROCAS

The number of images in the ALDRAM dataset is relatively small and there are no associated density labels so to apply supervised machine learning approaches we train our models on a separate data-set. In addition, by not using the ALDRAM data in the training process we maintain it as an independent test set.

The standard dose data-set we use for training is from the Predicting Risk Of Cancer At Screening (PROCAS) [14]. This data-set comprises mammograms from GE machines - the same as the ALDRAM data so we do not need to consider the issue of different imaging modalities due to machine variation.

Each image has two associated density estimates on a visual analogue scale (VAS) between 0 and 100 supplied by a pair of experienced mammogram readers [4]. The label is the average of these two scores. The pair of readers were pragmatically selected from a pool of 19 and there are differences in the distributions of density scores produced by individuals. Previous work has shown that differences in these distributions significantly affects the trained models [15]. To reduce this effect we use a subset of the PROCAS data which were all scored by the same pair of readers to avoid confounding the models due to variation in training labels.

This subset of the PROCAS data consists of images of the same size as the ALDRAM data (2, 294 × 1, 914 and 3, 062 × 2, 394). We use 15,290 mammograms which we partition into 12,287 training, 1,533 validation and 1,470 test images. The individual partitions include all the images available from one woman so there are no images from any one woman in different partitions.

Models trained on the standard dose PROCAS data-set might not produce accurate predictions on a low dose data-set. We therefore utilise software [16, 17] which can simulate low dose images using an input standard dose image. Using this software we generated simulated low dose images for each of the 15,290 mammograms to create a parallel data-set to train a low dose model on. The labels are the same as the standard dose equivalents.

## 3 Methodology

### 3.1 Image preparation

All images are pre-processed in the same way and are set to a final size of 1, 280 × 1, 024. LCC and LMLO images are flipped to the right side. We pad the smaller (2, 294 × 1, 914) sized CC images equally on top and bottom of the image to increase the size to 2, 995 and the smaller MLO images with 701 additional rows on the bottom of the image. The smaller CC and MLO images are padded on the left side with 480 new columns. The larger images are cropped to remove 34 rows. All images are now at size 2, 995 × 2, 394. The images are then resized using cubic interpolation to size 1, 280 × 1, 024. We clip the image intensities to 75% of their maximum and invert the pixel intensity. We perform histogram equalisation and normalise the images to between 0 and 1. This pre-processing approach is the same that produced the largest images in previous related work [8, 13]. There is a further normalisation step that takes place before training, and inference, which we will discuss in Section 3.2.

### 3.2 Model and training

We train two sets of models: one set on the PROCAS standard dose data and the second set on the PROCAS simulated low dose data. Our approach is identical for both the standard dose and simulated low dose data so that we minimise differences in outcomes due to reasons different from the level of radiation dose.

We use ResNet-18 [18] from the popular ResNet family of models and initialise the model using weights pre-trained on ImageNet [19]. The final fully connected layer that maps the representation to the 1,000 classes is removed and another fully connected layer is re-inserted with one output neuron. We do not apply any additional function to the output neuron.

As the output is continuous, we use the mean squared error (MSE) as our objective function and use the Adam [20] optimiser with hyper-parameters left as standard except for the learning rate which we use for model tuning. We train using a Nvidia V100 GPU with 16GB of RAM. The model parameters are saved at each epoch if the total validation error is lower than for any previous run. We utilise data augmentations with left-right flips and the additional of Gaussian noise, we also train with no data augmentation.

We considered the best performing two models from both the standard dose and the simulated low dose data-set. The final models are selected by their similarity in prediction to one another on the PROCAS test sets. This approach is taken, rather than selecting the models which perform best compared to the radiologist averaged scores because we are investigating the differences between the standard and low dose trained models.

### 3.3 Analysis approach

The aim of this work is to investigate the confidence we can place in the low dose predictions compared to standard dose equivalents, and if there are any approaches that can improve prediction similarity. To do so we investigate potential sources of variation which we specify below.

One source of variation in density prediction on mammograms is in variable predictions on CC and MLO images. Some studies [8] train separate models on the different views and others train models on both together [13] but both approaches tend to produce different density scores on the two views. Whether this is due to different information content in the views, model variability or other reasons is not clear.

Two different training runs of the same model will have different weights and produce different predictions. This may be due to alterations in hyper-parameters or for more stochastic reasons such as the assignment of training data to different batches. As we train two different models for the standard and low dose images, we have to consider how much of the variation in prediction is due to the model differences.

We have two different sized images (see Section 2.1) and they are consistent across the set of images, i.e. a low dose image will be originally the same size as a standard dose image. The two sizes of images have the same pixel size and in the pre-processing stage we altered the smaller image to match the larger image. However, there still might be variation in the model performance between the different image sizes.

The ALDRAM study was aimed at producing ratios of 10% between the low dose and standard dose radiation level. However, there was some variation in the actual ratio. As the low dose models were trained on simulated low dose data at 10% of the standard dose level, there may be effects on the quality of the prediction if the dose ratio of the ALDRAM data varies.

The ALDRAM dataset consists of younger women (see Figure 2) than those who consented to PROCAS. Younger women have generally denser breasts but there may also be other changes with age that cause the models to perform differently due to the different age distributions in the training set (PROCAS) from the testing set (ALDRAM).

Some automated models have shown poorer predictive accuracy at higher densities, at least partially because these are less well represented in training data [8, 21]. We might expect more variable predictions between the standard and low dose images at those higher densities due to poorer model (standard or low dose) performance. There may also be additional variation in performance between standard and low dose images at different densities.

Breast area may also have an impact on the accuracy of density predictions. In this paper we define breast area as the ratio of the non-background pixels to the background pixels. There may be effects from general model performance (both standard and low dose) and also specific to the low dose.

## 4 Results

### 4.1 Quality of the predictions on PROCAS

In Table 1 we show metrics on the PROCAS test set relating to the two pairs of models we selected. *Standard 1* and *Low 1* are the models used to make predictions on the ALDRAM data and the results of which make up most of our analysis. *Standard 2* and *Low 2* are the second best performing pair of models. The *Labels* refers to the averaged radiologist scores for the PROCAS data.

**Table 1.** Spearman rank correlation (*Rank Correlation*) and root mean squared error (*RMSE*) metrics on the PROCAS test set for the best performing two models on both standard and simulated low dose. *Standard 1* and *Sim. low 1* are the best performing pair of models when comparing the standard dose predictions to the simulated low dose predictions. Uncertainties were found via bootstrapping and reported at the 95% level. Results are shown both for the models against the labels (averaged radiologist scores) and against the other model predictions.

| Comparison |  | Rank Correlation | RMSE |
| --- | --- | --- | --- |
| Standard 1 | Labels | 0.860 (0.843-0.875) | 7.45 (7.10-7.82) |
| Sim. low 1 | Labels | 0.852 (0.834-0.868) | 7.71 (7.36-8.06) |
| Standard 2 | Labels | 0.857 (0.839-0.873) | 7.88 (7.51-8.25) |
| Sim. low 2 | Labels | 0.863 (0.845-0.879) | 7.44 (7.08-7.80) |
| Standard 1 | Sim. low 1 | 0.974 (0.971-0.977) | 2.65 (2.51-2.80) |
| Standard 2 | Sim. low 2 | 0.970 (0.965-0.974) | 3.36 (3.17-3.56) |
| Standard 1 | Sim. low 2 | 0.952 (0.946-0.957) | 3.85 (3.64-4.07) |
| Standard 2 | Sim. low 1 | 0.934 (0.926-0.941) | 3.95 (3.76-4.15) |

The best performing models with regard to the labels are not necessarily the best performing when we compare low dose and standard dose predictions. As we are interested in how the low dose predictions differ from the standard dose predictions on the ALDRAM data we selected the best performing model when considering the differences between standard and low dose predictions on the PROCAS test set. However, the performance of all the models, compared to the labels, is high. When considering Spearman’s rank correlation the predictions of all four models, when considered against the labels, all fall within the uncertainty bounds of each other. The aim of this paper is not to investigate how well the models perform against the labels, but in the relationship between the standard and low dose predictions because we do not have density labels for the ALDRAM data.

### 4.2 The effect of CC-MLO comparisons and multiple trained models

The predictions made by models on CC and MLO images can be variable for the same individual even though we use the same model to make predictions on both views. We show Spearman rank correlation coefficient and root mean squared errors (RMSE) of model predictions on standard and low doses for the RCC and RMLO in Table 2. The low dose differences are higher (but not statistically significantly) than the equivalents for the standard dose images, with a lower rank correlation and higher RMSE. Plots showing the values are in the appendix.

**Table 2.** Spearman rank correlation (*Rank Corr.*) and RMSE for the comparisons when considering RCC and RMLO prediction variation as discussed throughout Section 4.2. The sample uncertainty range, inside the brackets, is found by bootstrapping and reported at the 95% level.

| Related fig | Comparison1 | Comparison2 | Rank Corr. | RMSE |
| --- | --- | --- | --- | --- |
| 16 | RCC std. | RMLO std. | 0.962 (0.944-0.972) | 4.78 (4.15-5.38) |
| 16 | RCC low | RMLO low | 0.950 (0.918-0.969) | 4.66 (3.66-5.81) |
| 3 | RCC std. | RCC low | 0.964 (0.945-0.974) | 4.71 (4.15-5.27) |
| 3 | RMLO std. | RMLO low | 0.973 (0.959-0.979) | 4.35 (3.75-4.95) |
| None | RCC std. | RMLO low | 0.965 (0.948-0.973) | 5.23 (4.74-5.72) |
| None | RMLO std. | RCC low | 0.940 (0.907-0.960) | 6.08 (4.92-7.23) |
| 3 | Std. averaged | Low averaged | 0.980 (0.968-0.986) | 3.91 (3.44-4.36) |

In Figure 3 we show plots of the low dose predictions against the standard dose predictions for the RCC (left plot) and RMLO (middle plot) respectively. Related Spearman rank correlation and RMSE scores for these results are shown in Table 2. We also show, for completeness, comparison metrics from the alternate RCC, RMLO scores for standard and low dose in Table 2, i.e. for RCC low dose predictions compared to RMLO standard dose and vice-versa. The RMLO comparisons appears to show a more consistent result than the RCC but the result is not statistically significant. There is at least as much, and possibly more, variation between different views (RCC and RMLO) at the same dose, than there is between the same view but at different doses.

**Fig. 3.**
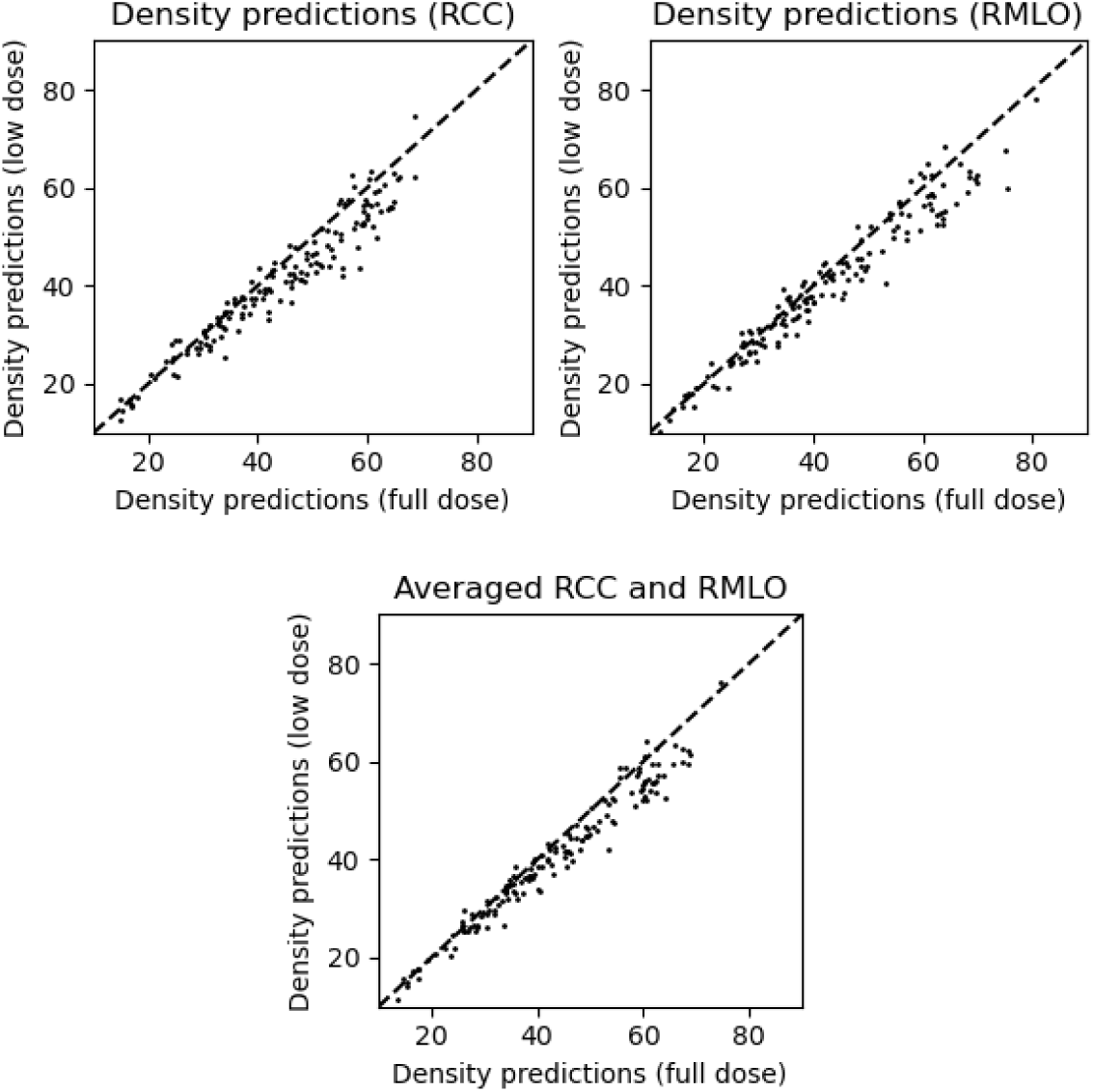
Plots of the low dose model predictions against standard dose model density predictions for the RCC view (top left) and RMLO view (top right). Also shown is a comparison of the standard and low dose predictions when averaging across the RCC and RMLO predictions (bottom). Metrics related to these plots are shown in Table 2.

We also show a plot of the low dose against standard dose predictions after averaging across the RCC and RMLO images (bottom plot of Figure 3). The metrics for the averaged scores are also in Table 2 named as *Std. averaged* and *Low averaged*. Averaging across the RCC and RMLO images produces greater similarity between the standard and low dose predictions.

Another potentially important decision is how to utilise the predictions of multiple trained models on the same data. We consider the effect of this model variation by performing two repeats of training for both the standard dose and low dose images - as discussed in Section 3.3. We show the effect of averaging across both the pairs of standard and low dose predictions on the absolute differences between the standard and low dose predictions in Figure 4. This averaging across models is called *Models combined* in the plot and the distribution of absolute differences is shifted towards smaller absolute differences - averaging the models produces more similar predictions.

**Fig. 4.**
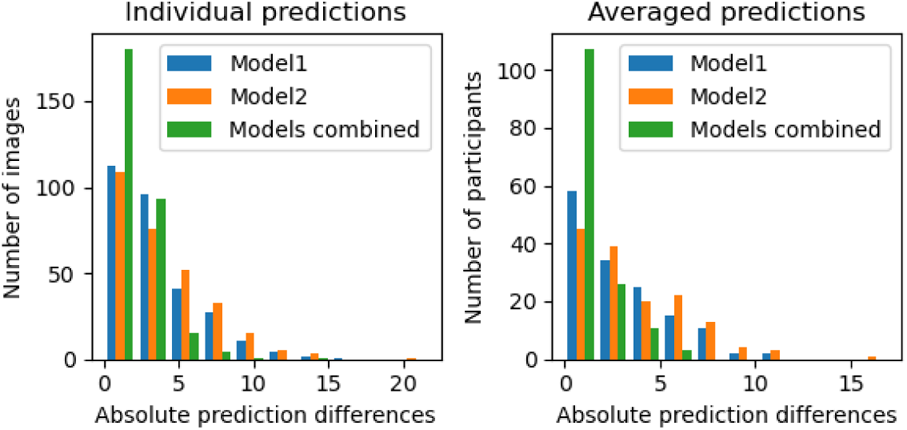
Absolute differences between standard and low dose predictions (in percentages) for two independent models (*Model 1* and *Model 2*) and the average of the two models (*Models combined*). Left) RCC and RMLO absolute prediction differences results separately. Right) Averaged across the RCC and RMLO predictions.

In Table 3 we further demonstrate the value of averaging across the models by showing the metrics between the low and standard dose for individual models and when pairs of models are averaged. When we average across both the standard and low dose pairs of models (labelled as *C*) the comparison metrics are statistically significantly improved.

**Table 3.**
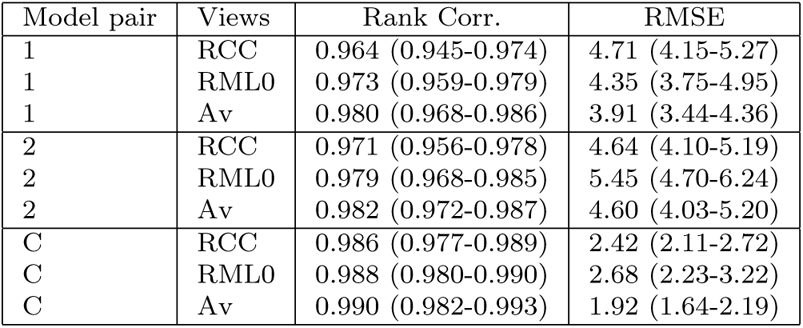
Spearman rank correlation and RMSE for the comparisons between standard and low dose images with different models and views. Model pair 1 produced the best performance on the PROCAS data and model pair 2 the second best. When the model predictions are averaged (labelled as *C*) the metrics improve significantly.

### 4.3 Factors of variation and uncertainty

We now consider factors that may have an impact on the accuracy of the low dose predictions. By investigating these factors, we can state whether they have an impact on the performance of the low dose models. These were all discussed in Section 3.3.

To investigate the relationship between prediction differences and age we show, in Figure 5, plots of the absolute differences in prediction for three different age ranges: younger than 40, between 40 and 42 (inclusive) and older than 42. The left plot shows a boxplot of the absolute prediction differences for the averaged views. In the right plot we show the mean average absolute prediction difference per age range along with uncertainty at the 95% level found by bootstrapping for both the individual (*Ind.*) and averaged (*Av.*) predictions. There is no statistically significant difference in performance in the different age ranges. Additional results for the quality of prediction with age are shown in the appendix.

**Fig. 5.**
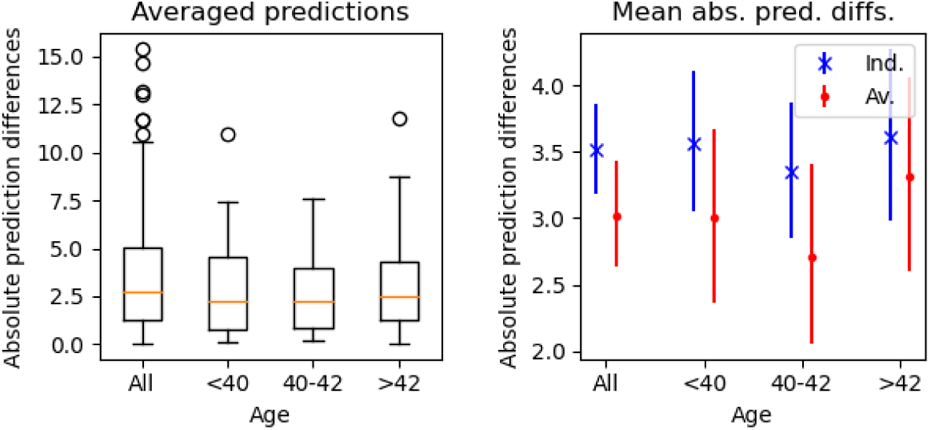
Variation in differences with age found via separating women into the age bins. Left) the boxplot shows the median, range and any outliers for the absolute prediction differences on averaged views. Right) the mean absolute prediction differences on low and standard dose images for individual (*Ind.*) and averaged (*Av.*) predictions. The uncertainty bounds are found via bootstrapping and are reported at the 95% level.

The magnitude of the density score is a potentially important factor for the quality of the low dose predictions. In Figure 6 we show the differences in predictions (standard dose minus low dose predictions) against the average of the predictions across low and standard dose - how the difference in prediction changes as the density of the breasts increases. We see an increase in prediction variation as the density increases. This is not just an increase in variation in prediction difference but the standard dose models are making higher density predictions compared to the low dose models.

**Fig. 6.**
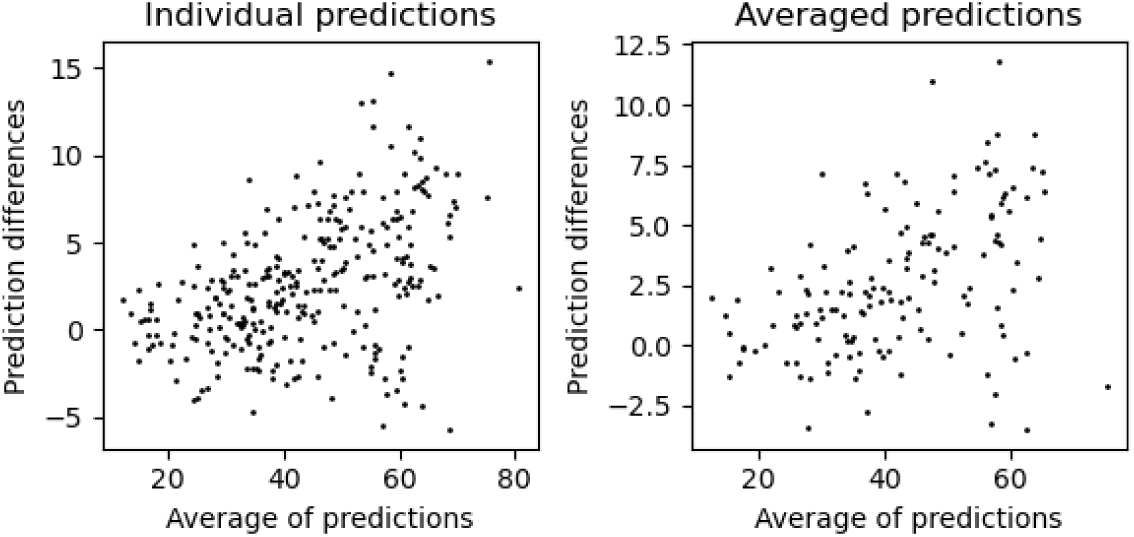
How the differences in predictions between standard and low dose models varies with density, as defined by averaging across the low and standard dose predictions. Left) individual views, both RCC and RMLO plotted separately. Right) averaged across RCC and RMLO.

We further emphasise the importance of the level of density to the confidence we can place in the low dose model in Figure 7 where we separate the images into density bins of *<* 36, 36 *−* 51 (inclusive) and *>* 51. We see an increase in absolute prediction differences at the higher densities. The left plot shows a box-plot for the averaged predictions and the right plot shows averaged absolute prediction differences with uncertainty bounds calculated via bootstrapping and shown at the 95% level. The three plots show the same increase in prediction difference at higher density.

**Fig. 7.**
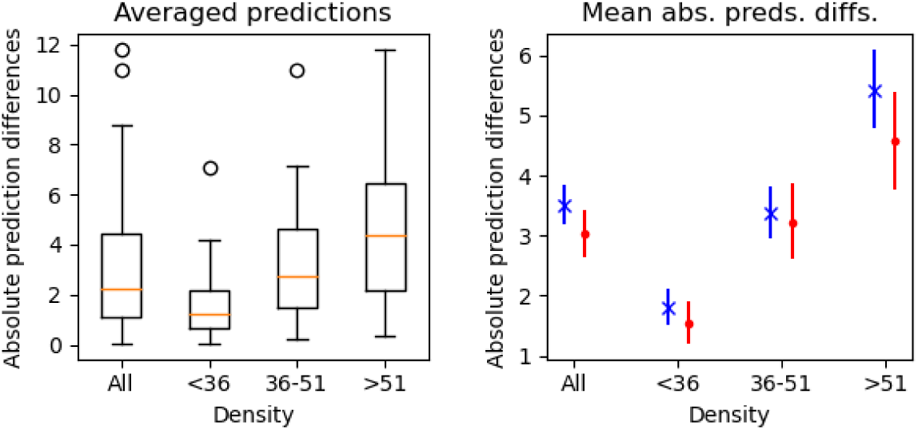
Plots of the absolute differences between standard and low dose averaged (across RCC and RMLO) predictions for different density ranges. The data is found from separating the results into density bins of *<* 36, 36 *−* 51 (inclusive) and *>* 51. averaged predictions. Left) boxplots of the absolute density differences in each bin. Right) plots with mean average absolute prediction differences within each bin and uncertainty found by bootstrapping at the 95% level.

While the variation in the dose ratios is not large (see Figure 2), there might be some differences in performance due to these small variations. In Figure 8 we demonstrate that there is no evidence for any difference at different doses - the Spearman rank coefficient (0.002) shows no relationship. The left plot shows boxplots when we split the dose ratios into two groups with lower dose ratios and higher dose ratios, with the dividing point at 0.0954. The right plot shows the same split but with the mean average absolute differences with uncertainty generated via bootstrapping.

**Fig. 8.**
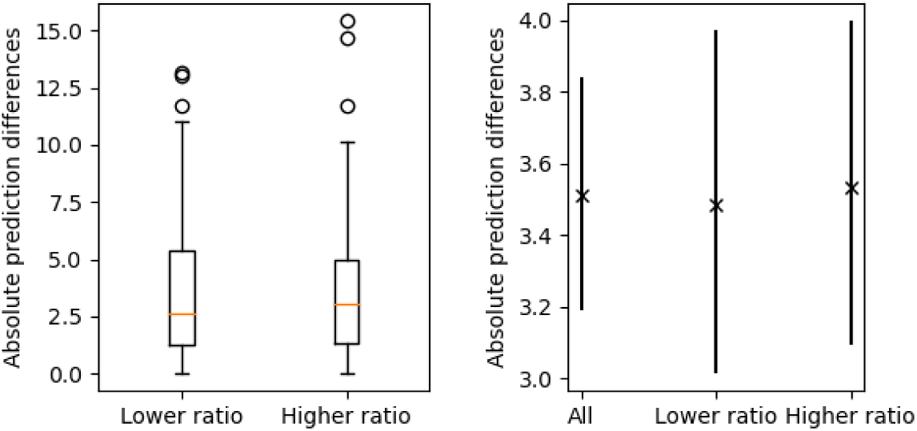
Plots showing the lack of a relationship between the absolute prediction difference between standard dose and low dose density scores and the exposure ratio. All results are for individual images, not averaged. Left) Boxplots of the absolute prediction differences for those pairs of images with an exposure ratio below and above 0.0954 respectively. Right) Mean average absolute prediction differences for the different groups with the uncertainty found via bootstrapping.

The ALDRAM data is produced at two image sizes and in Figure 9 we demonstrate that this factor may correlate with differences in performance. The left plot shows histograms of the distributions of the absolute differences for the two sizes for the results when averaged across RCC and RMLO which are noted by “1” (3, 062× 2, 394) and “2” (2, 294× 1, 912). We see that at size 2 there appears to be significantly more images with large differences. The right plot shows the mean absolute prediction differences with uncertainty produced by bootstrapping. There is a difference in prediction performance between the two differently sized images.

**Fig. 9.**
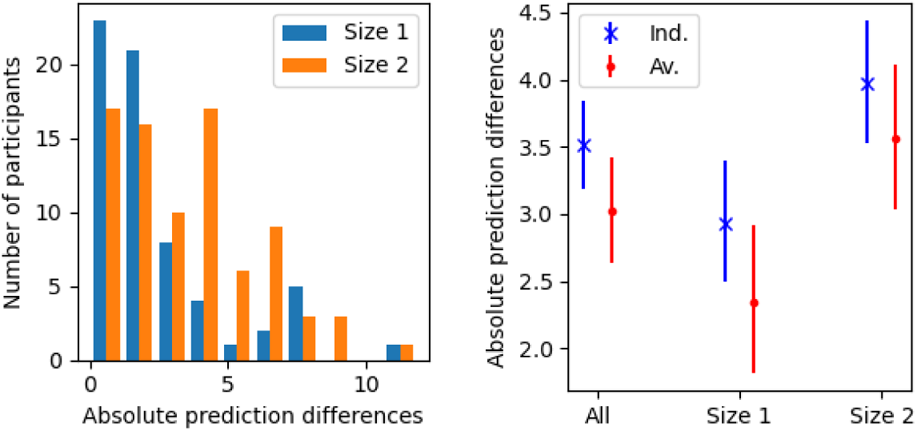
Absolute differences between standard and low dose predictions by image size. Left) histogram showing the distribution of absolute differences for the averaged predictions. Right) the mean absolute prediction differences with uncertainties for the different sizes and for individual (*Ind.*) and averaged (*Av.*) predictions.

The area of the breast, which we define here as the ratio of pixels that are not background to background, can have an effect on the results. In Figure 10 we show the distribution of breast area in the left plot. The area of the breast, compared to the whole image, has a peak at around 0.25 with a relatively small number of images which occupy over half the image. The right plot shows the averaged prediction differences against the breast area, which is also averaged. There is a significant negative correlation between the differences in prediction and the breast area, with a Spearman rank correlation of −0.48. We show the results on the CC and MLO images combined but there is no difference in conclusion if we consider them separately with a Spearman rank coefficient of approximately −0.40 between prediction differences and the breast area for both individual sets of predictions.

**Fig. 10.**
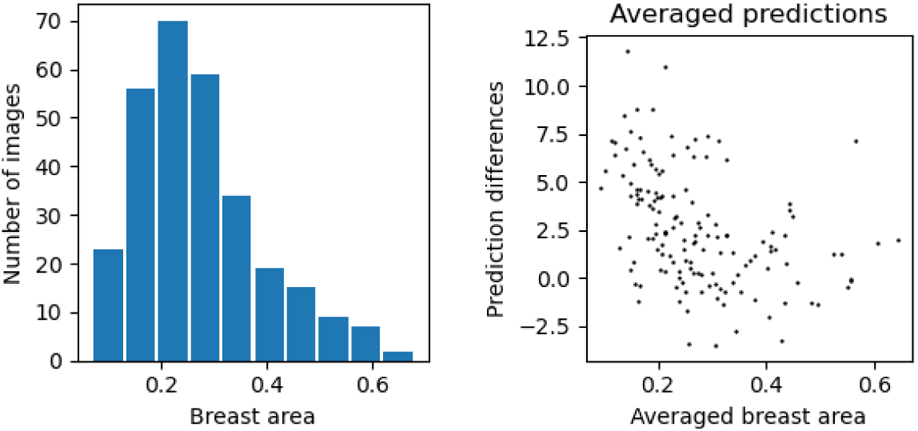
Left) A histogram showing the distribution of breast areas for all the images, defined as the ratio of the breast tissue to background. Right) The differences in prediction against the breast area when averaging across the CC and MLO predictions. The breast area is also averaged across the RCC and RMLO images.

In Figure 11 we show results when we bin the images into four quartiles based upon the breast size. The left plot is a boxplot for the averaged prediction differences. The right plot shows the average prediction differences with uncertainty found via bootstrapping and shown at the 95% confidence level. We are not considering absolute results in these plots, so they underestimate the level of difference of prediction performance. The low dose model shows a substantial under-prediction of the breast density compared to the standard dose model for small breast areas. There is little difference at larger breast areas. Additional results are shown in the appendix.

**Fig. 11.**
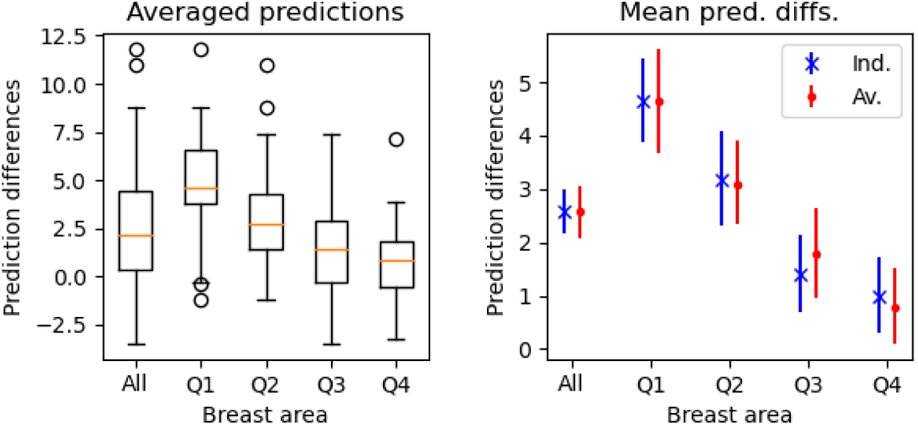
Differences in prediction by quartiles of breast area. Left) Boxplot of the prediction differences for averaged predictions. Right) Average prediction differences with uncertainty found via bootstrapping and reporting at the 95% level.

This variation in performance between the standard and low dose images from breast area is likely to be the reason for the differences in performance on different image sizes. In Figure 12 we demonstrate this by showing the relationship between the breast area in the mammogram and the image size. The left plot shows the distributions of the breast area for the two different image sizes. The right plot shows the average with uncertainty of the breast area at the two different image sizes. The larger image sizes show breasts with significantly larger area which likely accounts for the variation in performance on the different image sizes.

**Fig. 12.**
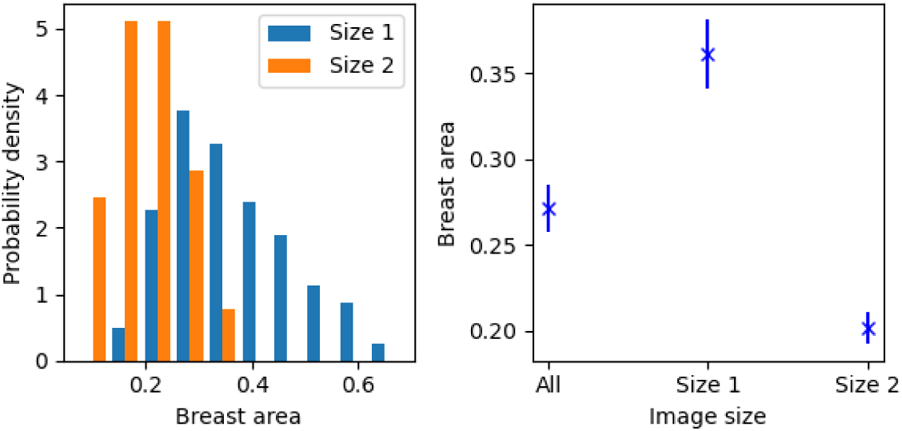
The relationship between image size and breast area. Left) Histograms showing the distribution of the breast area for the two image sizes (*Size 1* and *Size 2*). Right) Average breast area with uncertainty found via bootstrapping and reporting at the 95% level for the two image sizes as well as them both combined.

Breast area and mammographic density are also related. In Figure 13 we show the relationship between the density predictions and the breast area for the standard (left plot) and low (right plot) dose images. There is a negative correlation between predicted density and breast area with a Spearman rank coefficient value of −0.51 and −0.44 for the standard and low dose images respectively. Both of these factors - the density and the breast area - may individually contribute to greater uncertainty about the quality of the low dose predictions but they are also correlated.

**Fig. 13.**
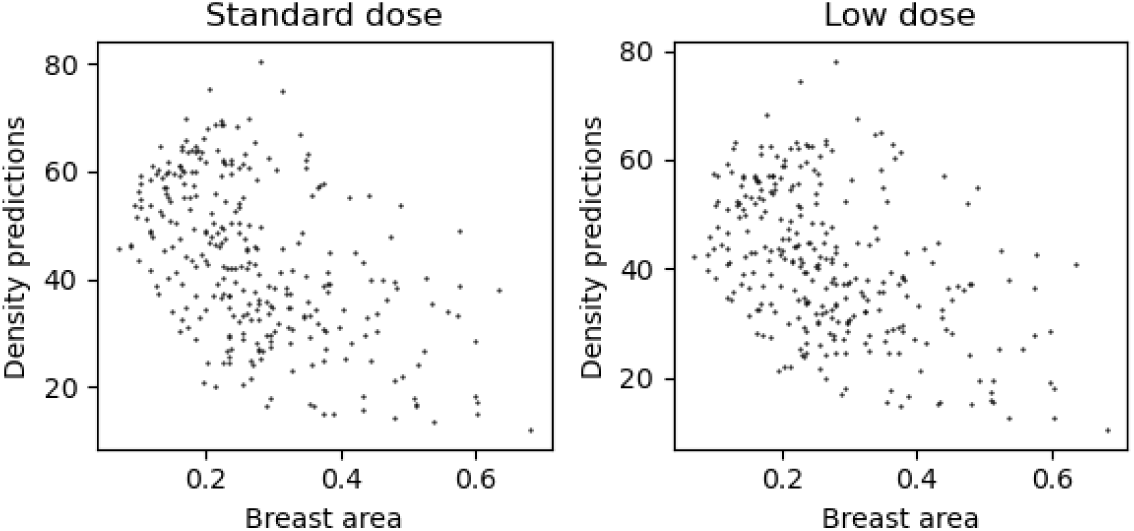
The relationship between breast area and predicted density. Left) Standard dose images. Right) Low dose images.

In Figure 14 we show what information can be gained by looking at the differences between RCC and RMLO low dose predictions and the difference between standard dose and low dose. The plots show the absolute differences for ranges of the low dose RCC and RMLO differences. The left plot shows all the results as a boxplot and the right plot shows the averages for each range with uncertainty from bootstrapping and showing the 95% uncertainty bounds. When the gap between the RCC and RMLO for the low dose is high we are also more likely to see a larger difference between the low and standard dose predictions. However, from these results, this is only really true at relatively large differences between the low dose CC-MLO predictions with there being little relationship if we exclude the most extreme differences.

**Fig. 14.**
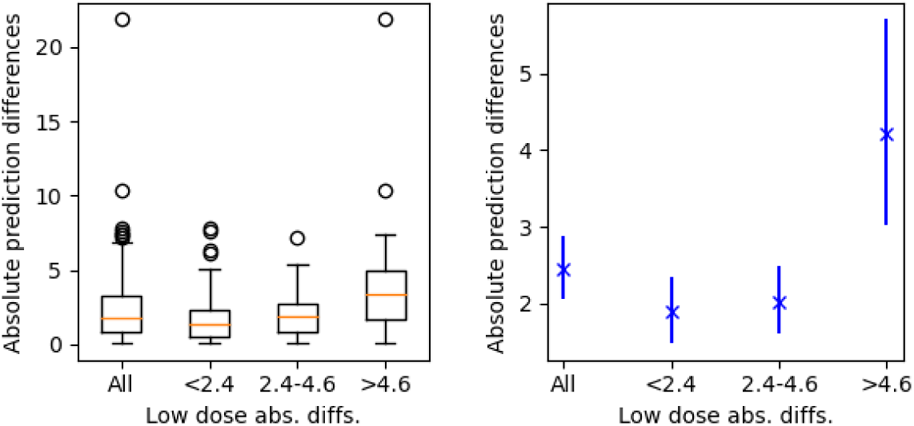
Plots of the absolute differences between the averaged standard dose and averaged low dose predictions against the absolute differences between the RCC and RMLO predictions of the low dose images. Left) Boxplots after splitting the data into bins for the absolute differences between RCC and RMLO. Right) The average and uncertainty of the absolute differences for differences in the RCC versus RMLO results.

In Figure 15 we show what information can be gained by examining the differences between two separately trained low dose prediction models. The plots show the absolute differences for ranges of the low dose repeated model prediction differences. The left plot displays boxplots showing all the results and the right shows the averages for each range with uncertainty from bootstrapping and showing the 95% uncertainty bounds. With large differences in predictions between the pair of low dose predictions we also see larger differences between the low dose and standard dose predictions.

**Fig. 15.**
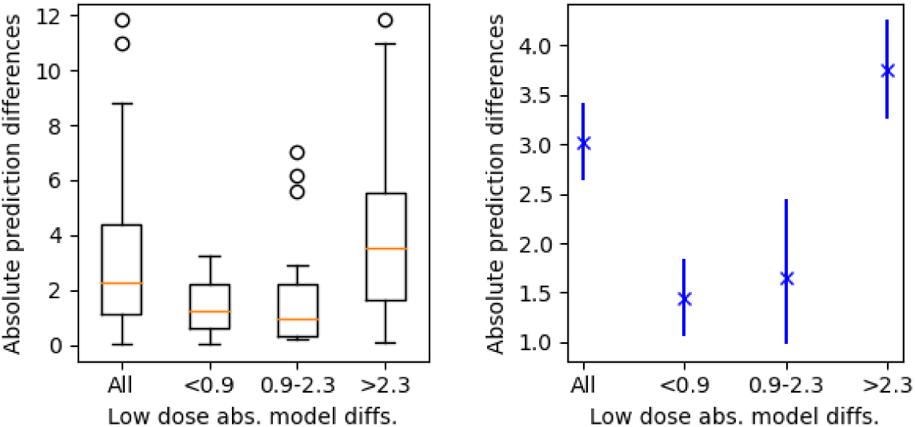
Plots of the absolute differences between the averaged standard dose and averaged low dose predictions against the absolute differences between a pair of trained model of the low dose images. Left) boxplots after splitting the data into bins for the model absolute differences. Right) the average and uncertainty of the absolute standard-low differences for differences in averaged model predictions.

We summarise, in qualitative terms, the effect of the variables on the performance of the low dose models in Table 4. We show the variable, which figures and tables they link to (including those in the appendix) and give a description of what effect the variable has on the performance of the model on low dose images.

**Table 4.** Qualitative summary of our findings. We show the variable, which figures and tables they link to and give a description of what effect the variable has on the performance of the model on low dose images

| Variable | Evidence | Comments |
| --- | --- | --- |
| Age | Figs 18,5 | No effect. |
| Density | Figs 6,7 | Substantial effect with poorer performance at high densities. |
| Dose ratio | Figs 8 | No effect. |
| Image size | Figs 9 | Poorer performance on smaller image sizes, likely due to distribution of breast area. |
| Breast area | Figs 10, 11.<br>Table 6 | Substantial effect with poorer performance on smaller breast area. |
| CC-MLO differences | Figs 14 | Poorer performance when large difference in CC-MLO predictions. |
| Model differences | Figs 15 | Greater differences in predictions between models may imply greater variation between low and standard dose predictions. But likely dependent on pair of models compared. |

The final consideration is about potential outliers. In Table 5 we show how many standard dose - low dose image pairs that have an absolute difference greater than a set of numbers from 2 to 12. So for CC the number of image pairs (standard dose and low dose) that have an absolute difference greater than 2 is 99 and the fraction of all the images is 0.67. The averaged scores have mostly fewer at each level than either the CC or the MLO - averaging tends to reduce the low-standard dose prediction differences. We also see that only very small numbers have differences larger than 10% points. From these results we do not see any notable outliers - there is a fairly smooth progression down to the larger differences. Figures for these outlier results are shown in the appendix.

**Table 5.** Outlier table. On the left side of the table is the absolute difference value. The *Number* is the number of pairs of images (low and standard dose) which have a larger absolute difference between predictions that the absolute difference number. So for the CC view 99 images out of 147 have a larger absolute difference than 2. The *Fraction* is the fraction of images with absolute differences larger than the specified value.

| Abs. diffs. | CC |  | MLO |  | Averaged |  |
| --- | --- | --- | --- | --- | --- | --- |
|  | Number | Fraction | Number | Fraction | Number | Fraction |
| 2 | 99 | 0.67 | 89 | 0.61 | 82 | 0.56 |
| 4 | 51 | 0.35 | 43 | 0.29 | 46 | 0.31 |
| 6 | 31 | 0.21 | 24 | 0.16 | 23 | 0.16 |
| 8 | 12 | 0.08 | 11 | 0.07 | 5 | 0.03 |
| 10 | 5 | 0.03 | 4 | 0.03 | 2 | 0.01 |
| 11 | 4 | 0.03 | 3 | 0.02 | 1 | 0.01 |
| 12 | 2 | 0.01 | 2 | 0.01 | 0 | 0.00 |

## 5 Discussion

The use of low radiation dose mammography enables the assessment of mammographic density in younger women to allow for improved prediction of individual breast cancer risk prior to routine breast screening. This may lead to earlier diagnosis and improved health outcomes. Strategies for breast cancer risk assessment of women between 30 and 39 are currently being investigated [11].

Deep learning methods have been shown to make accurate mammographic density estimates on standard dose mammograms [8]. Recent work has also shown that the same type of models can make predictions on low dose mammography that are similar to their standard dose counterparts [12, 13]. This offers the potential for a fully automated method to be able to estimate accurate density, and therefore risk, predictions on younger women.

Deep learning models can produce highly impressive performance but they mostly lack transparency; we do not fully understand why many models make the predictions they do. In addition, these types of models can be brittle - giving substantial changes in predictions with only small changes in the inputs. It is always important to understand uncertainty in any prediction, but these two factors make it particularly important when using deep learning. In this paper we studied predictions of mammographic density made by deep learning algorithms on a paired standard and low dose mammography dataset to provide evidence of the level of confidence we can place in the low dose predictions.

We used a model drawn from one of the most commonly used model families, ResNet, and trained two sets of models: one for standard dose and one for low dose images. The standard dose images were taken from the PROCAS mammography data-set and the low dose image models were trained on simulated low dose images created from the PROCAS data-set. The predictions made on the ALDRAM standard dose data-set act as our ground truth.

We demonstrate that combining the predictions made on CC and MLO images reduces the variation between standard and low dose images but may be at the cost of some lost information and compression of the distribution. Our results suggest that there may be as much difference in density predictions between CC and MLO images of the same dose as the same view with different doses, but this would need to be confirmed using a larger dataset.

Generally combining multiple models improves performance. By using our two best performing models (for both the standard and low dose images) on the validation set and averaging the test set predictions we find more similar predictions between the standard and low dose models than from a single model.

We investigated the factors that are correlated with variation in low and standard dose predictions and summarise the results in Table 4. We see no relationship between performance and either age or the dose ratio. Age in particular might have had a significant impact on performance as our models are trained on women generally older than the women in the ALDRAM dataset.

We can also consider the differences in prediction on CC and MLO images as a marker for poorer performance. This might be an expected result but it is useful in that CC and MLO images are usually produced and it is worthwhile knowing that if the gap between the predictions on the two views for the low dose images is substantial then the gap between low dose and standard dose predictions is also likely to be large. It is also not inevitable - it might have been the case than divergence in prediction between CC and MLO images on the low dose images would be matched in the standard dose images meaning the gap between the standard and low dose predictions would be unrelated to the gap between CC and MLO.

The difference in performance between two high performing (on the simulated low dose validation set) models on individual images may point towards increased variation between low and standard dose images. Our results do show this but it is somewhat variable with the choice of pairs of models to compare. We should be less confident if the differences in pairs of models on one low dose image is high and it may indicate less trust can be placed in that prediction but it is likely to be model dependent.

We demonstrated a link between the performance on the low dose images and the image size but also showed that this is most likely due to the compressed breast area rather than anything to do with differential model performance on the different sized images.

There is a substantial correlation between model performance and breast area with poorer performance on smaller breasts. However, model performance is also worse on denser breasts. Smaller breasts also tend to be denser so this is a heavily confounded. We can conclude that the model predictions on both smaller and denser breasts requires some correction.

We also demonstrate the lack of significant outliers in our data-set. There is a reasonable distribution of differences without any images being outside this distribution.

The results we have shown demonstrate that we can make highly similar predictions on low dose compared to standard dose mammograms. This is especially true if we average across both CC and MLO views and use predictions from more than one model on the mammograms. However, there are also systematic differences in prediction which should be investigated with low dose predictions being generally lower than the standard dose counterparts. There are also issues with the relationship between similarity in prediction (between standard and low dose) and both breast area and the density level. Finally, we considered the interdependence between image size and breast area and also between breast size and density but did not do so for the other factors. It may be the case than some of the other factors are also correlated. A full causal model [22] may be necessary to extract the interrelated relationships that effect the uncertainty level we have on the low dose predictions, potentially with calculated corrections for low dose density results to take forward into risk prediction algorithms.

## A Additional results

In Figure 16 we show plots of model predictions on the ALDRAM standard dose (left) and low dose (right) images of RMLO against RCC.

**Fig. 16.**
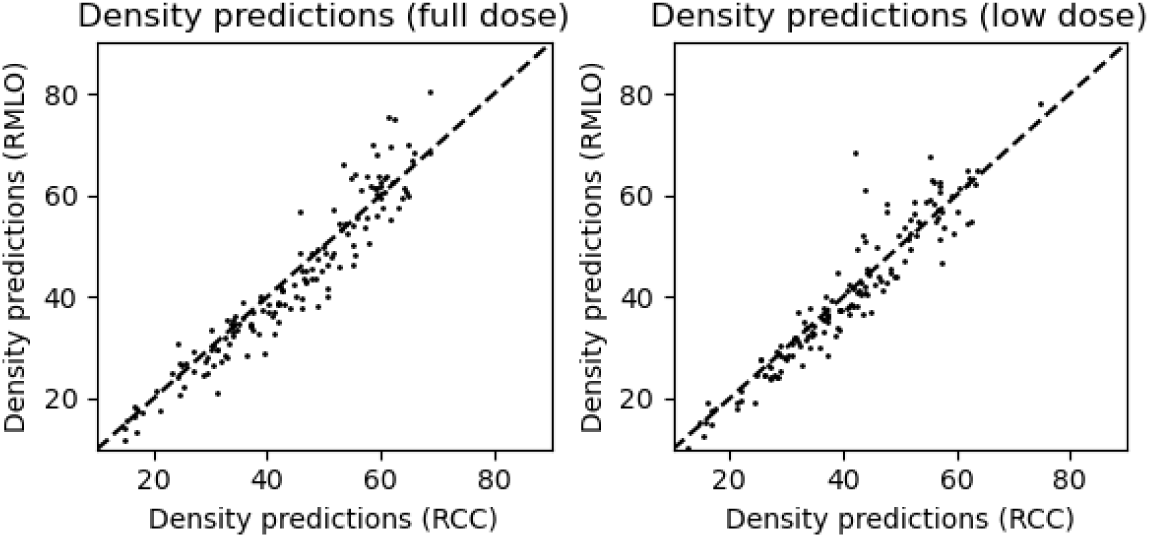
Plots of the model density predictions on RMLO against RCC for the standard dose (left) and low dose (right) images. Metrics related to these plots are shown in Table 2.

In Figure 17 we show the distribution of the predictions for the RCC, RMLO and when averaging across RCC and RMLO. Averaging across the views should improve performance if the differences are due to noise. However, if the views contain different information averaging them might reduce prediction quality. Although it is modest, we may be able to see this effect in Figure 17 where the predictions are shifted somewhat towards the middle of the distribution by the averaging effect. The averaging process slightly compresses the distribution which may result in some information loss.

**Fig. 17.**
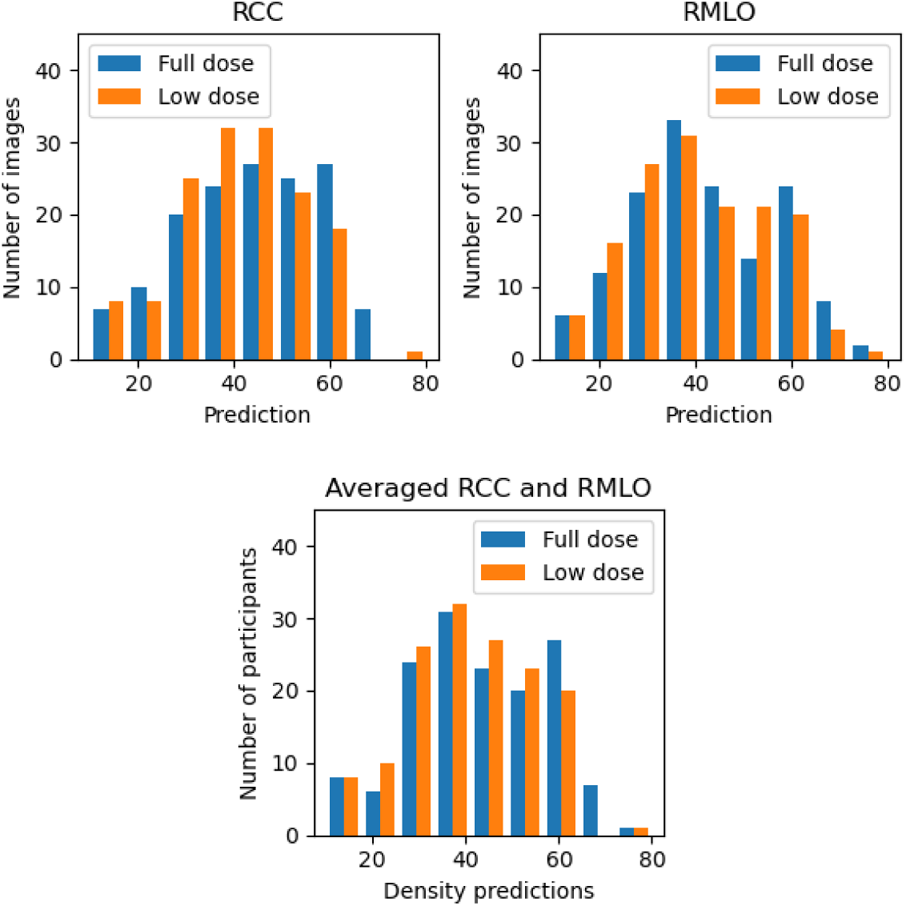
Histograms showing the distribution of the density predictions for the standard dose and low dose models. Top left) Histograms for the RCC predictions. Top right) Histograms for the RMLO predictions. Bottom) Histograms when we have averaged across the RCC and RMLO predictions.

To test the impact of age we display the prediction differences between the standard and low dose models against age in Figure 18. The left plot shows prediction differences for individual views (both RCC and RMLO differences are included in the plot) and the right plot shows the differences for the averaged views (averaging between RCC and RMLO predictions). We see no relationship with age with a Spearman rank coefficient of 0.06 for the individual predictions and 0.08 for the averaged predictions.

**Fig. 18.**
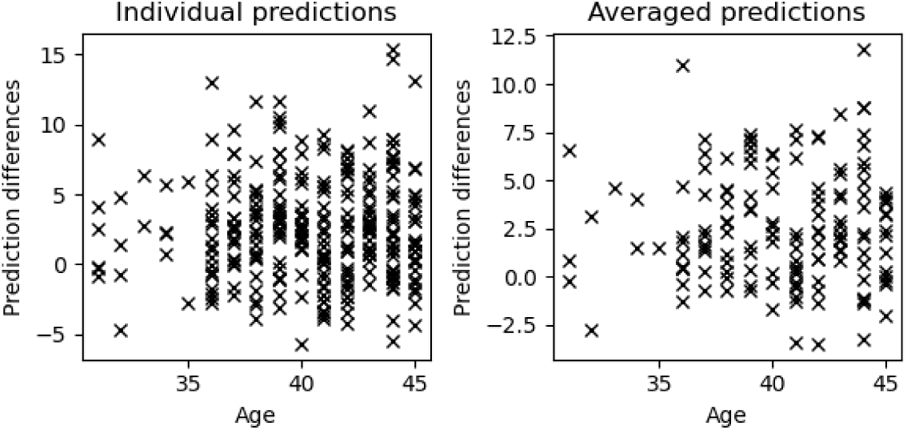
Prediction differences between standard dose and low dose images against the age of the woman. Left) individual views, both RCC and RMLO plotted separately. Right) averaged across RCC and RMLO.

Figure 19 shows direct plots of the absolute prediction differences between standard and low dose images against the exposure ratio, with no correlation between the two.

**Fig. 19.**
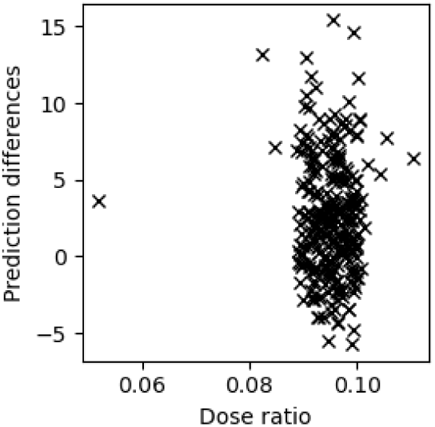
Plot showing the lack of a relationship between the absolute prediction difference between standard dose and low dose density scores and the exposure ratio. The absolute prediction difference plotted against the exposure ratio for individual images.

In Figure 20 we show boxplots of the individual and averaged predictions for image size 1 and size 2, denoted by “1” and “2” respectively.

**Fig. 20.**
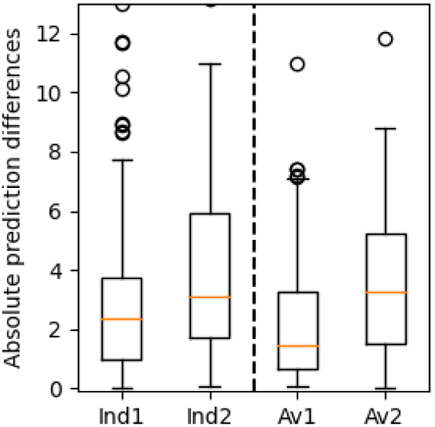
Absolute differences between standard and low dose predictions by image size. Boxplots of the individual and averaged predictions for size 1 and size 2, denoted by “1” and “2” respectively.

To summarise the variation in predictions between the low and standard dose for the different breast sizes we show the Spearman rank correlation and RMSE in Table 6. The results show substantial differences in performance with the similarity between the low and standard dose predictions being higher at the larger breast sizes.

**Table 6.** Spearman rank correlation and RMSE for the comparisons between standard and low dose images with different breast sizes.

| Quartile | View | Rank Corr. | RMSE |
| --- | --- | --- | --- |
| All | RCC | 0.964 (0.945 – 0.974) | 4.71 (4.14 – 5.28) |
|  | RMLO | 0.973 (0.958 – 0.979) | 4.35 (3.75 – 4.95) |
|  | Av | 0.98 (0.968 – 0.986) | 3.91 (3.45 – 4.37) |
| 1 | RCC | 0.878 (0.719 – 0.951) | 5.77 (4.4 – 7.13) |
|  | RMLO | 0.878 (0.717 – 0.947) | 6.48 (5.24 – 7.71) |
|  | Av | 0.882 (0.697 – 0.961) | 5.47 (4.57 – 6.35) |
| 2 | RCC | 0.971 (0.926 – 0.987) | 5.02 (4.16 – 5.82) |
|  | RMLO | 0.961 (0.895 – 0.986) | 4.15 (3.05 – 5.25) |
|  | Av | 0.976 (0.924 – 0.992) | 4.0 (3.08 – 4.91) |
| 3 | RCC | 0.964 (0.916 – 0.982) | 4.16 (3.08 – 5.2) |
|  | RMLO | 0.951 (0.884 – 0.977) | 3.55 (2.69 – 4.39) |
|  | Av | 0.973 (0.931 – 0.987) | 3.33 (2.55 – 4.04) |
| 4 | RCC | 0.962 (0.902 – 0.98) | 3.7 (2.47 – 4.91) |
|  | RMLO | 0.956 (0.859 – 0.989) | 2.38 (1.86 – 2.89) |
|  | Av | 0.985 (0.949 – 0.995) | 2.19 (1.54 – 2.91) |

In Figure 21 we show histograms of the absolute prediction differences between the standard dose and low dose image predictions for RCC (top left), RMLO (top right) and averaged across the views (bottom).

**Fig. 21.**
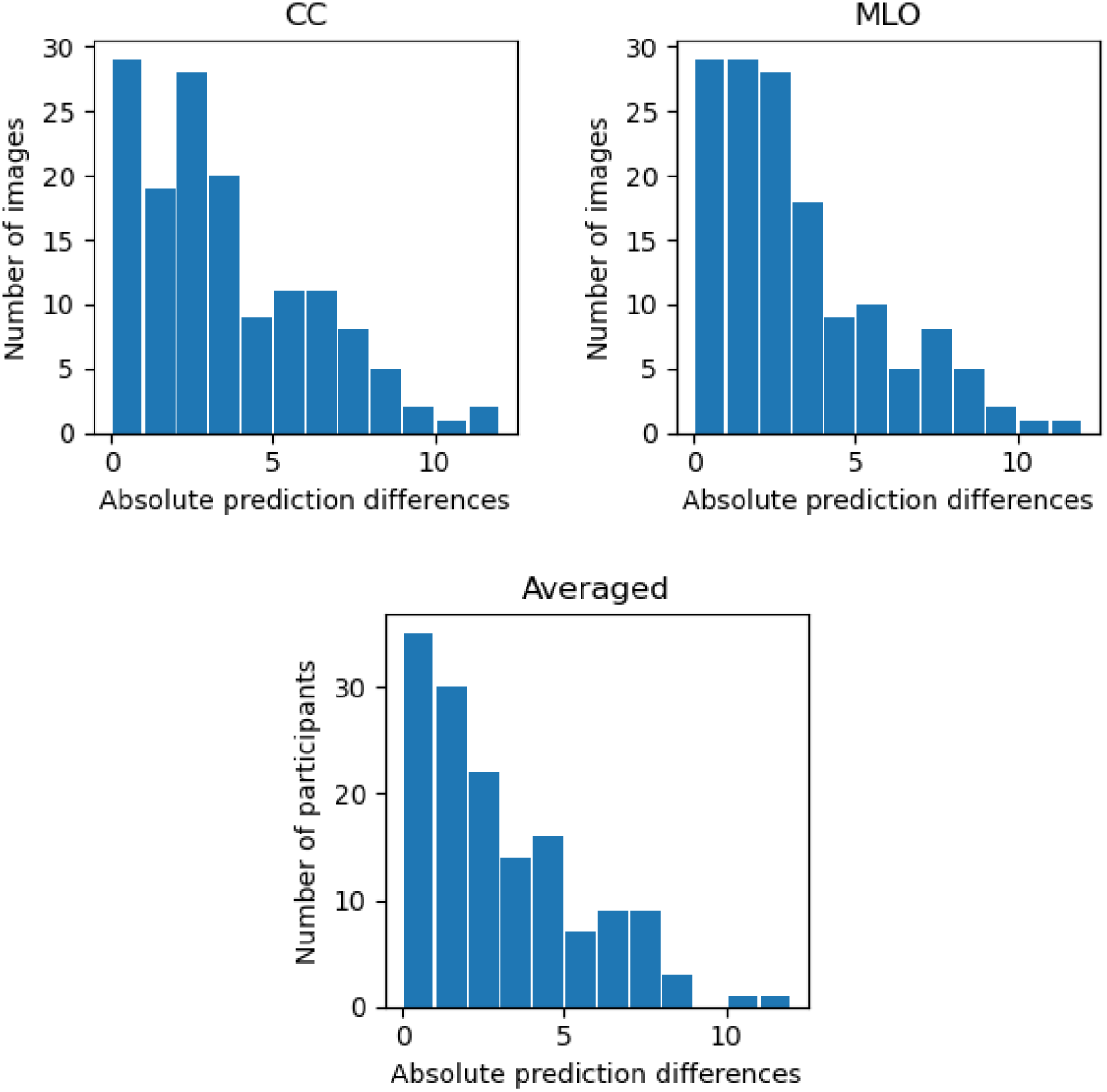
Histograms showing the frequency of the absolute predictions differences between the standard and low dose images. Top left) averaged differences. Top right) RCC differences. Bottom) RMLO differences.

## Data Availability

The data is not available to researchers outside of the University of Manchester.

